# Serum Neurofilament Light is Elevated in COVID-19 Positive Adults in the ICU and is Associated with Co-Morbid Cardiovascular Disease, Neurological Complications, and Acuity of Illness

**DOI:** 10.1101/2021.04.28.21256277

**Authors:** Meredith Hay, Lee Ryan, Matthew Huentelman, John Konhilas, Christina Hoyer-Kimura, Thomas G. Beach, Geidy E. Serrano, Eric M. Reiman, Kaj Blennow, Zetterberg Henrik, Sairam Parthasarathy

## Abstract

In critically ill COVID-19 patients, the risk of long-term neurological consequences is just beginning to be appreciated. While recent studies have identified that there is an increase in structural injury to the nervous system in critically ill COVID-19 patients, there is little known about the relationship of COVID-19 neurological damage to the systemic inflammatory diseases also observed in COVID-19 patients. The purpose of this pilot observational study was to examine the relationships between serum neurofilament light protein (NfL, a measure of neuronal injury) and co-morbid cardiovascular disease (CVD) and neurological complications in COVID-19 positive patients admitted to the intensive care unit (ICU). In this observational study of one-hundred patients who were admitted to the ICU in Tucson, Arizona between April and August 2020, 89 were positive for COVID-19 (COVID-pos) and 11 were COVID-negative (COVID-neg). A healthy control group (n=8) was examined for comparison. The primary outcomes and measures were subject demographics, serum NfL, presence and extent of CVD, diabetes, sequential organ failure assessment score (SOFA), presence of neurological complications, and blood chemistry panel data. COVID-pos patients in the ICU had significantly higher mean levels of Nfl (229.6±163 pg/ml) compared to COVID-neg ICU patients (19.3±5.6 pg/ml), Welch’s t-test, p =.01 and healthy controls (12.3±3.1 pg/ml), Welch’s t-test p =.005. Levels of Nfl in COVID-pos ICU patients were significantly higher in patients with concomitant CVD and diabetes (n=35, log Nfl 1.6±.09), and correlated with higher SOFA scores (r=.5, p =.001). These findings suggest that in severe COVID-19 disease, the central neuronal and axonal damage in these patients may be driven, in part, by the level of systemic cardiovascular disease and peripheral inflammation. Understanding the contributions of systemic inflammatory disease to central neurological degeneration in these COVID-19 survivors will be important to the design of interventional therapies to prevent long-term neurological and cognitive dysfunction.

## Introduction

Neurological complications following SARS-CoV-2 infection and been reported by a number of investigators (1), (2),(3). In addition, there is overlap of the risk factors in patients with severe COVID-19 and patients at risk for Alzheimer’s Disease Related Dementias (ADRD) and vascular contributions to cognitive impairment and dementia (VCID). These include age, hypertension, diabetes, cardiac disease, hypercholesterolemia, and pulmonary disease. Recent studies have shown that of ICU admitted COVID-19 patients in France, 84% showed some level of neurological impairment during their hospital admission (4) and there is a high prevalence of neurological involvement in critically ill patients suffering from COVID-19 (5), (6),(7),(8). It has been suggested that SARS-CoV-2 results in damage to the CNS damage via a surge of systemic inflammatory cytokines called Cytokine Storm Syndrome (CSS) (9),(10).

Neurofilament light protein (Nfl) is one of the 3 primary neurofilament isotypes that have been shown to increase in both the cerebrospinal fluid (CSF) and blood in the presence of axonal damage and neurodegeneration (11). Levels of serum Nfl have been found to be elevated in subjects with a number of neurological degenerative diseases (12),(13),(14), (15), as well as those with acute conditions such as hypoxic brain injury (16), cardiac and related surgeries (17), (18), and traumatic brain injury (19). Recent studies have found increased levels of Nfl in COVID-19 patients (20), (21),(22) but the effects of concomitant systemic inflammatory disease such as cardiac disease or diabetes on neuronal injury as measured by NfL in COVID-19 patients has not been previously reported. In the present study we examined the relationship between levels of serum Nfl in COVID-19 positive ICU patients and the presence of cardiovascular disease, diabetes, and acute neurological complications.

## Materials and Methods

### Study Population

One-hundred patients admitted to the ICU in Tucson, Arizona, USA, were included in this study. Of these, 89 patients were positive for COVID-19 with an average age of 60.8. Of these 89, full clinical data was available on 50 patients. All 50 of the COVID-19 positive (COVID-pos) ICU patients were mechanically ventilated. We compared the COVID-19-pos ICU Nfl data to 11 patients who were admitted to the ICU but were COVID-negative (COVID-neg), with an average age of 65.1. Of these 11, full clinical data were available on 5 patients. Of these five patients, four were mechanically ventilated and one was on a high flow nasal cannula system.

Blood samples for the Nfl assay were obtained on the day of admission to the ICU. The Nfl assay on samples from the ICU patients was performed in the Clinical Neurochemistry Laboratory at the Sahlgrenska University Hospital using the Single molecule array (Simoa) NF-light Advantage and HD-X Analyzer, (Quanterix, Billerica, MA). A single batch of reagents was used; intra-assay coefficients of variation were below 6.8% for all analytes. The Nfl assay on samples from the healthy control cohort were performed by PBL Assay Science, New Jersey, USA, and analyzed using the same Simoa NF-light Advantage kit on an HD-X Analyzer, as per the manufacturer’s instruction (Quanterix, Billerica, MA).

Upon admission, patient’s medical histories and clinical data including primary admission diagnosis, their SOFA score (the SOFA score (23) is derived from scores from six organ systems ranging from 0 (no organ dysfunction) to 4 (severe organ dysfunction) and the individual organ scores are then summed to a total score between 0 and 24), the Glasgow Coma Score (assessed without sedation), Systemic Inflammatory Response Syndrome (SIRS) evaluation, sepsis evaluation, pulmonary function including ventilation rates and volumes, days intubated, renal function, liver function, full chemistry laboratory panel, ICU-related complications, duration of stay in ICU, duration of hospital stay and in-hospital death. Cardiovascular disease (CVD) secondary diagnosis upon admission included the presence of heart failure, hypertension (defined as greater than 140-159/90-99 mm Hg), coronary artery disease, valvular disease, arrhythmia, hyperlipidemia, previous MI, obesity, and smoking history. The CVD score was derived by assigning one point for each of the above cardiovascular related diagnoses and then summing all for the total CVD score. Neurological complications (neurocx) were assessed upon hospital admission and during ICU stay. Levels of ICU delirium were assessed twice daily while in the hospital by trained research personnel using the Richmond Agitation-Sedation Scale (RASS) and Confusion Assessment Method-ICU CAM-ICU (24).

### Statistical Analysis

Experimental values are expressed as mean ± SE unless otherwise indicated. Comparison of the Nfl levels and log Nfl levels between the 2 ICU groups and the control were analyzed with Welches’ t-test with significance determined at a p value < .05. The Welches’ t-test was chosen due to the unequal variances and unequal sample sizes between the COVID-pos and COVID-neg groups. One COVID-pos patient Nfl value of 14,555 pg/ml was considered an outlier and not included in the statistical analysis. Comparison of log Nfl levels across subgroups were analyzed with Kruskal-Wallis ANOVA with p < .05 considered significant. The Kruskal-Wallis ANOVA was chosen due to the non-normal distribution of Nfl levels across groups. Associations between log Nfl levels and other co-morbidities and clinical laboratory variables were analyzed using Pearson correlation and a p value < .05 indicated statistical significance. Linear regression was used to generate a best-fit line. Data were analyzed using GraphPad Prism 8.0.

### Human Subject Protection and Protocol Approvals

This study and use of patient samples was approved by the University of Arizona Institutional Review Board (IRB# 1410545697). Participant informed consent was provided either directly by the patient or, if the participant was incapacitated, from the patient’s legally authorized representative.

## Results

### Demographics

All ICU patients had either a rt-PCR confirmed positive or negative result for SARS-CoV-2 infection. The three groups analyzed included 89 COVID-pos ICU patients, 11 COVID-neg ICU patients, and 8 healthy controls. Both ICU patient groups were recruited from Tucson, AZ between April-August 2020. The healthy adult participants were recruited independently from emergency care personnel in the hospital. Age deciles for each group are detailed in **Table 1**. The average age of the COVID-pos ICU patients was 60.8, the average age of the COVID-neg ICU patients was 65.1, and the average age of the healthy controls was 51.7.

**Table 1.** Age Deciles of Cohort Groups.

| Age Decile |  |  |  |  |  |  |  |  |  |
| --- | --- | --- | --- | --- | --- | --- | --- | --- | --- |
| ***Participant Cohort*** | **n** | **Avg Age** | **20-29** | **30-39** | **40-49** | **50-59** | **60-69** | **70-79** | **80-89** |
| COVID-19-pos ICU patients | 89 | 60.8 | 3 | 1 | 5 | 11 | 15 | 13 | 2 |
| COVID-19-neg ICU patients | 11 | 65.1 |  |  | 1 | 1 |  | 4 |  |
| Control healthy adults | 8 | 51.7 | 1 | 1 | 1 | 3 | 1 |  | 1 |

Demographic and initial admission clinical data for both COVID-pos and COVID-neg ICU patients are detailed in **Table 2**. Of the 89 COVID-pos ICU patients, we had full demographic and clinical data on 50 patients. All 50 of the COVID-19 positive (COVID-pos) ICU patients were mechanically ventilated. Of these 50 COVID-pos patients, 39 (43.8 percent) did not survive. Of the 11 COVID-neg ICU patients who we had plasma samples from, full demographic and clinical data were available in 5 patients. Of these five patients, four were mechanically ventilated and one was on high flow nasal cannula system. 4 out of the 11 (36.3 percent) did not survive. Levels of critical illness as measured by the total sequential organ failure assessment score (SOFA) were not different between the COVID-pos and COVID-neg groups (9.5±0.4 vs 10.8±2.2, respectively). Cardiovascular disease (CVD) was noted in 35 of the 50 COVID-pos patients and some exhibited more than one type of CVD. This included 29 with hypertension (defined as 140-159/90-99 mm Hg), 10 with hyperlipidemia, 10 with a previous/old MI, 6 with coronary artery disease, 4 with arrhythmia, 2 with heart failure, 1 with valve disease, 3 with obesity and 6 with a smoking history.

**Table 2.**
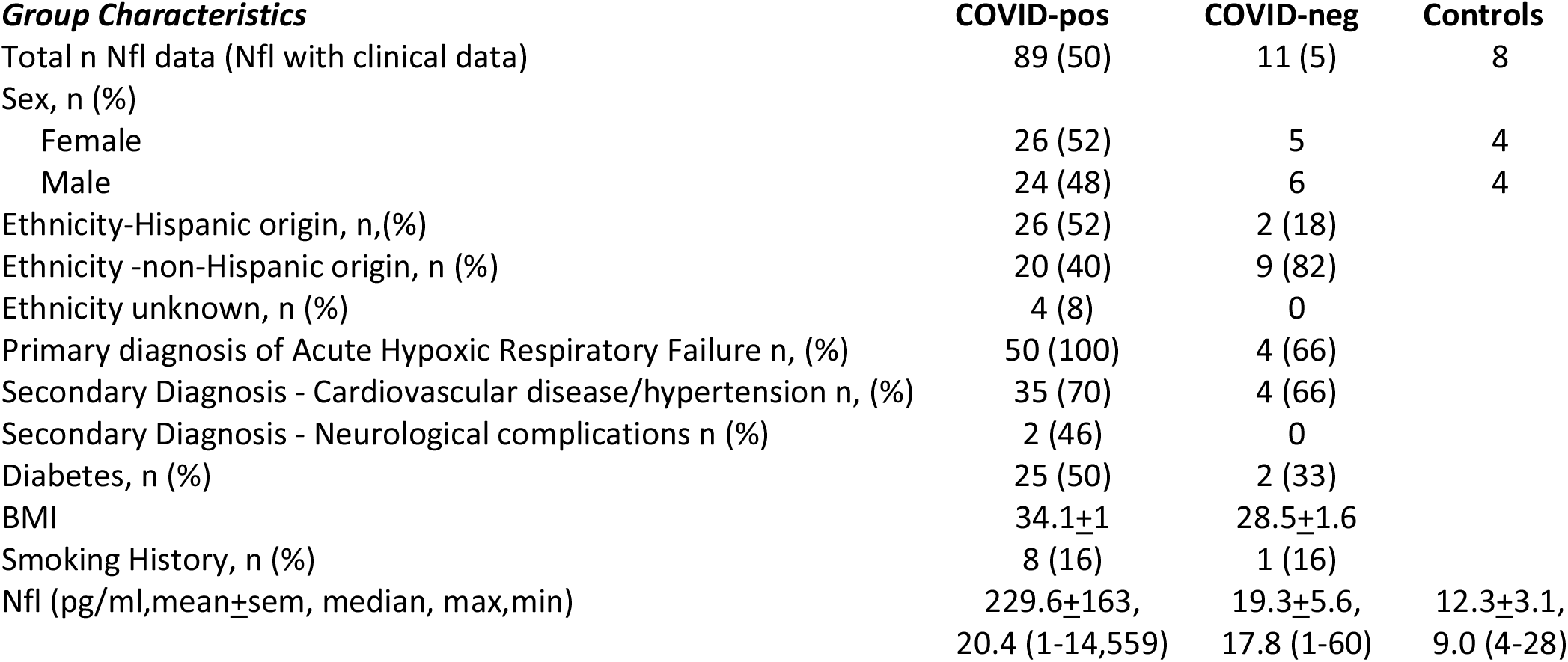
Demographic, clinical and Nfl levels.

Neurological complications (neurocx) were assessed upon hospital admission and during ICU stay. Of the 50 COVID-pos patients, 5 were diagnosed while in the ICU with acute encephalopathy, 5 with ICU delirium, 4 with sleep apnea, 2 with seizure, 5 with depression, one with cerebral edema, 2 with stroke, and one with facial droop. None of the 5 COVID-neg ICU patients had any noted neurological complications.

### Nfl Biomarker

In COVID-pos patients in the ICU, Nfl was significantly higher than those observed in COVID-neg ICU patients and healthy control (**Figure 1A**). COVID-pos patients had a mean plasma Nfl of 229.6±163.9 sem pg/ml, median of 20.4 pg/ml (min 1.0 - max 14,555) vs. COVID-neg patients with a mean plasma Nfl level of 19.3±5.6 sem pg/ml, median of 17.8 pg/ml (min 1.0 - max 60.2), (95% CI 8.18 to 86.78, two-tailed Welch’s t-test, p = .01). The 8 healthy controls had mean Nfl value of 12.3±3.1 sem pg/ml, median 9.0 pg/ml (min 3.7- max 27.9) and were significantly different from COVID-pos (95% CI 16.27 to 92.71, two-tailed Welches’ t-test, p =.005).

**Figure 1.**
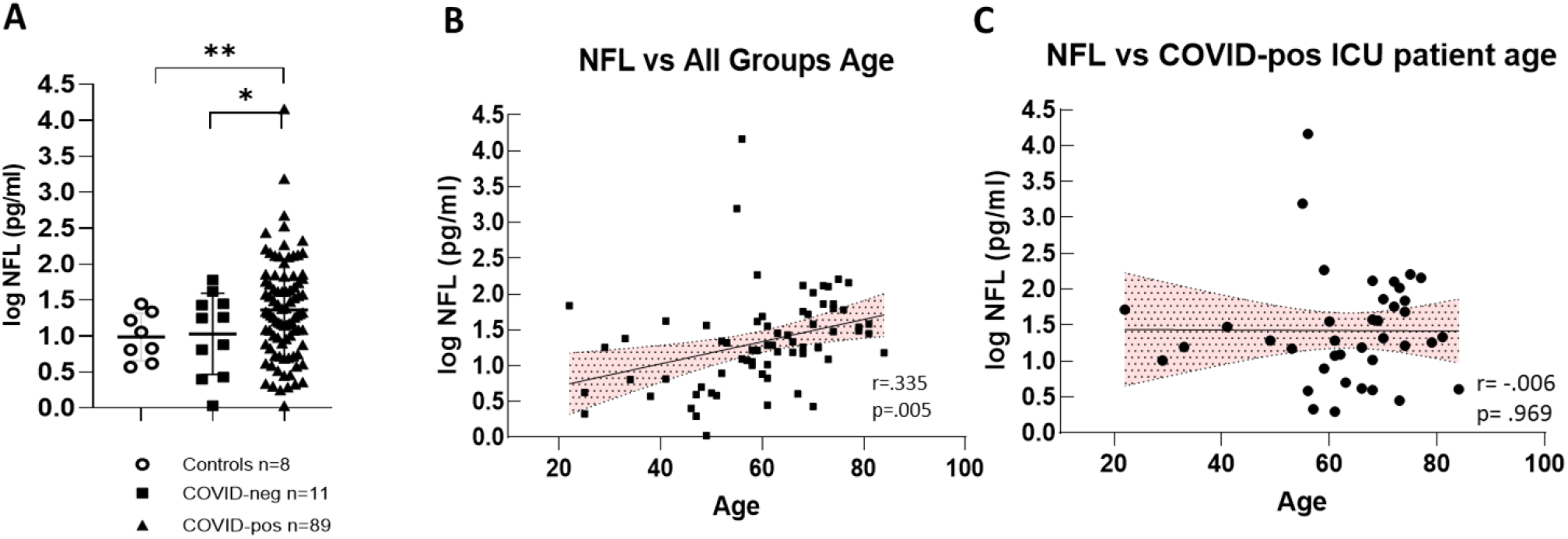
**(A)** Illustrates log Nfl values from COVID-pos, COVID-neg ICU patients and healthy controls. COVID-pos mean plasma Nfl levels were 66.85±18.9 sem pg/ml, median of 19.8 pg/ml (max 1555-min 1.0 vs. COVID-neg, mean plasma Nfl levels were 19.3±5.6 sem pg/ml, median of 17.8 pg/ml (max 60.2-min 1.0), (95% CI 8.18 to 86.78, two-tailed Welch’s t-test, p =, 01). The 8 healthy controls had mean Nfl levels of 12.3±3.1 sem pg/ml, median 9.0 pg/ml (max 27.9-min 3.7) and were significantly different from COVID-pos (95% CI 16.27 to 92.71, two-tailed Welches’ t-test, p =.005). **(B)** Log Nfl levels and age from all the groups analyzed together show that log Nfl levels were significantly correlated to age in the combined groups, (r = .335, p =.005, Pearson). **(C)** COVID-pos ICU patient Nfl levels and age were analyzed separately and there was no significant correlation to age (r = .006, p = .969, Pearson).

To determine if the levels of Nfl in groups were related to subject age, we first performed Pearson correlation analysis of all log Nfl values compared to all group ages (**Figure 1B**). In this case, the levels of Nfl were significantly positively correlated with age, (r = .335, p =.005, Pearson), as has been previously reported (12),(13). However, when we performed correlation analysis of Nfl values compared to age in COVID-pos ICU patients alone (**Figure 1C)**, there was no significant correlation of Nfl levels with the age of the COVID-pos patient (r = .006, p = .96).

### Nfl and Cardiovascular Disease

There is a clear and well established association between the presence of CVD and the risk of cognitive impairment, neurodegenerative disease and vascular dementia (26),(27),(28), (29),(30),(31), (32),(33). In the present study, we tested the hypothesis that levels of Nfl in COVID-pos ICU patients would be associated with the presence of CVD and diabetes. As seen in **Figure 2F**, of the 50 COVID-pos patients, 38 had at least one form of CVD with hypertension being the most prevalent with 29/50 COVID-pos patients having a diagnosis of hypertension. **Figure 2D** illustrates the distribution and variance of CVD plotted against the level of Nfl for each cardiovascular disease category. As seen in **Figure 2A**, log Nfl levels were significantly higher in COVID-pos patients with CVD (mean 1.68±.09 sem, median 1.57, min 1.0-max 4.1 log pg/ml) compared to those with no known CVD (mean 1.0±0.16 sem, median 0.89, min 0.2 – max 3.1 log pg/ml), (95% CI .27 to 1.0, two-tailed, Welches’s t-test, p = .001). Likewise, the levels of Nfl in COVID-pos patients were significantly higher in those who had diabetes (mean 1.9±0.1, median 1.8 min 1.3 – max 4.1) as compared to those that did not have diabetes (mean 0.9±.06 sem, median 1.0, min 0.3 – max 1.3) (95% CI 0.70 to 1.2, two-tailed, Welch’s t-test, p<.0001). (**Figure 2B**). **Figure 2C** compares mean log Nfl levels between COVID-neg patients (n=11), COVID-pos patients with no CVD and no neurocx (n=9), COVID-pos with CVD and no neurocx (n=18) and those with both CVD and neurocx (n=23). While the highest levels of Nfl were seen in COVID-pos patients with both CVD and neurocx there was no significant difference in log Nfl levels in COVID-pos with CVD and no neurocx and those with both. The log Nfl levels in COVID-pos patients were positively correlated with the composite CVD Score (**Figure 2E**), (r = 0.34, p = .001, Pearson) suggesting that the Nfl levels in these COVID-pos patients are related to the level of CVD.

**Figure 2.**
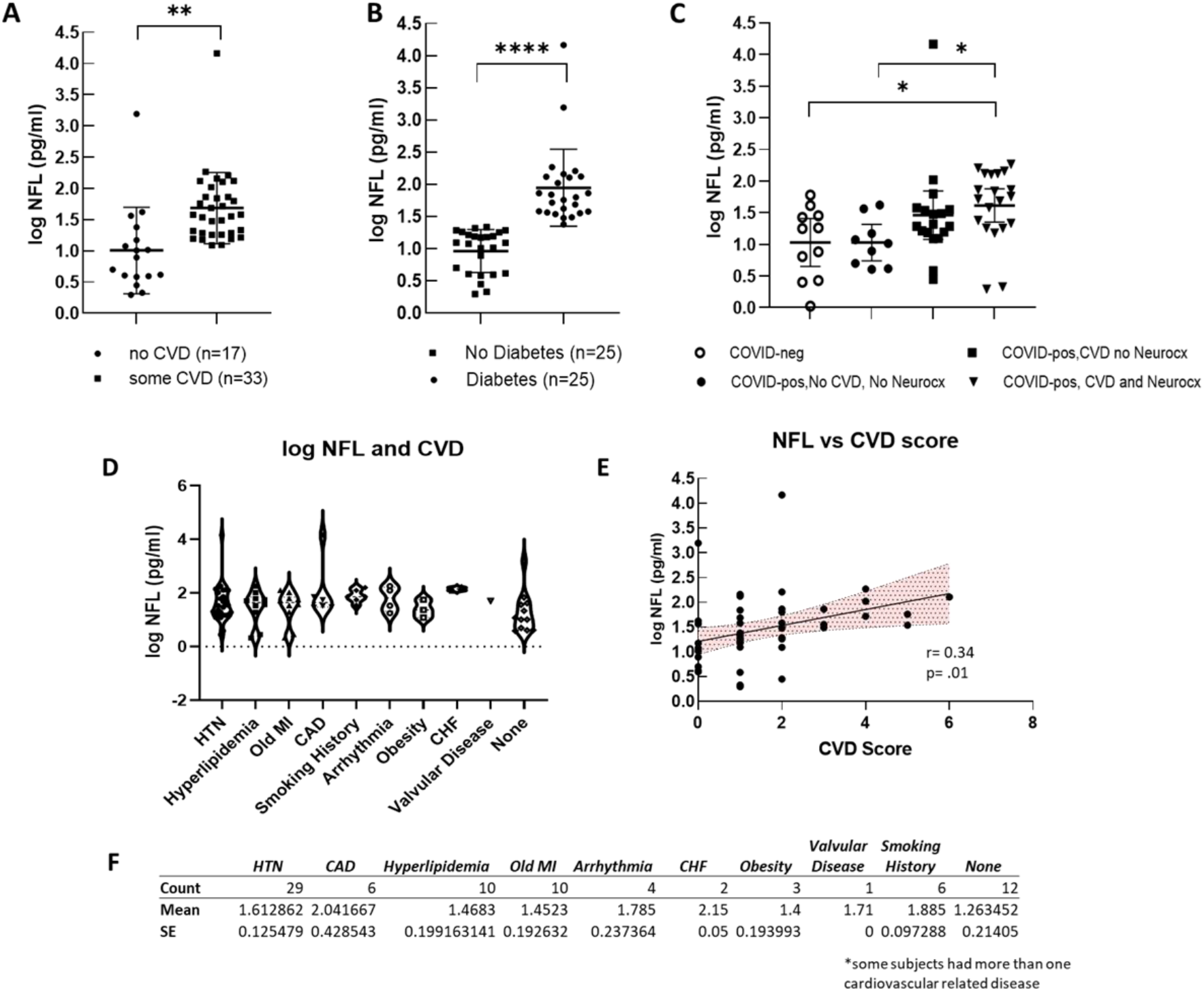
**(A)** Compares log Nfl values from COVID-pos patients with either some level of CVD or none. The log Nfl levels were significantly higher in COVID-pos patients with some level of CVD (median 1.57, min 1.0-max 4.1 log pg/ml) compared to those with no known CVD (median 0.89, min 0.2 – max 3.1 log pg/ml), (95% CI .27 to 1.0, two-tailed, Welches’s t-test, p = .001). Nfl levels were significantly higher in COVID-pos patients with diabetes (median 1.8 min 1.3 – max 4.1) as compared to those that did not have diabetes (median 1.0, min 0.3 – max 1.3) (95% CI 0.70 to 1.2, two-tailed, Welch’s t-test, p<.0001). **(C)** Compares log Nfl levels between COVID-neg (n=11), COVID-pos with no CVD and no Neurocx (n=9), COVID-pos with CVD and no Neurocx and COVID-pos (n=18) with both CVD and Neurocx (n=23). While the highest levels of Nfl were seen in COVID-pos with both CVD and Neurocx there was no significant difference in log Nfl levels in COVID-pos with CVD and no Neurocx and those with both. **(D)** Illustrates the distribution and variance of cardiovascular disease in COVID-pos patients plotted against the level of Nfl for each cardiovascular disease category. **(E)** shows the correlation between the levels of Nfl and the CVD score in COVID-pos patients. The levels of NfL were significantly positively correlated with the level of the CVD score (r = 0.34, p = .001, Pearson). **(F)** Is a table comparing the mean ± sem of log Nfl levels in each of the categories of cardiovascular disease.

### Neurofilament Light and Neurological Complications

To determine if the levels of Nfl were related to the presence of neurocx, we compared Nfl levels in COVID-pos ICU patients in those that had at least one reported neurocx to those that had none. Of the 50 COVID-pos ICU patients, 23 exhibited at least one or more neurocx and 27 had no neurocx. Of these 23, all also had at least one or more noted diagnosis of CVD. The types and distributions of neurocx in the COVID-pos patients are seen in **Figure 3A and 3C**. None of the 5 COVID-neg ICU patients exhibited any neurocx. **Figure 3B** illustrates significant differences in mean log Nfl levels in COVID-pos with neurocx (mean 1.6±0.1, median 1.7, min 0.29 - max 3.1, n=23) compared to COVID-pos with no observed neurocx (mean 1.2±.07, median 1.2, min 0.4 – max 2.0, n=27), 95% CI 0.7 to .73, two-tailed Welch’s t-test, p =.01).

**Figure 3.**
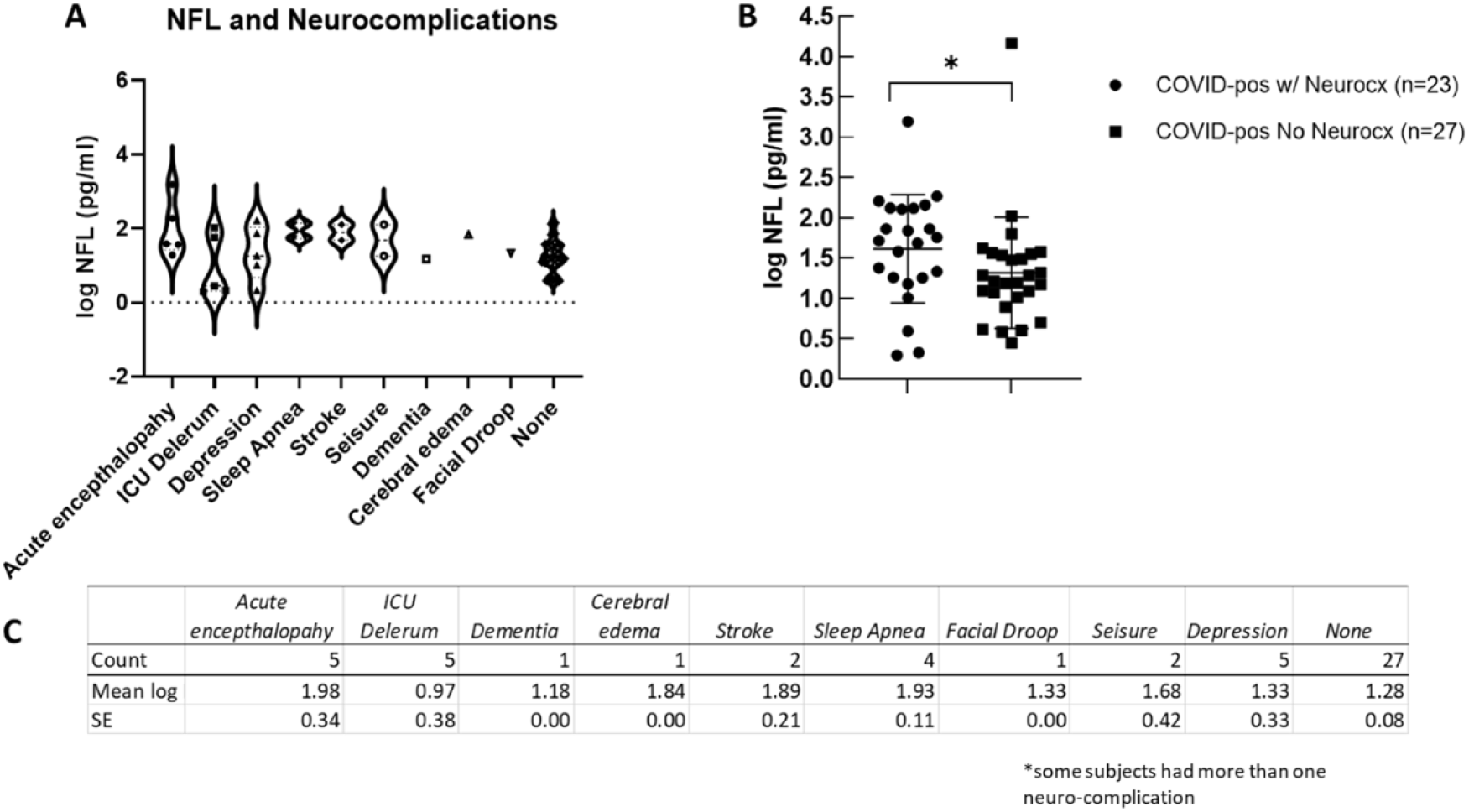
**(A)** Illustrates the distribution and variance of neurological complications in COVID-pos patients plotted against the level of log Nfl for each neurological complication category. **(B)** Illustrates a significant difference of log Nfl levels in COVID-pos patients with neurological complications (1.6±0.1, median 1.7, min 0.29-max 3.1, n=23) to COVID-pos patients with no observed neurological complications (1.2+.07, median 1.2, min 0.4 – max 2.0, n=27), 95% CI 0.7 to .73, two-tailed Welch’s t-test, p =.01). **(C)** Is a table comparing the mean ± sem of log Nfl levels in each of the categories of neurological complication.

### Nfl Levels: ICU Status and Clinical Blood Panel Markers

All 50 of the COVID-pos patients exhibited sepsis and multiple organ system failure and systemic inflammatory response syndrome (SIRS), all of which have been associated with post ICU cognitive impairment (24),(34). We evaluated the association between COVID-pos Nfl levels and the ICU clinical status and sequential organ failure assessment core (SOFA). As seen in **Figure 4A**, in COVID-pos patients, the levels of Nfl were correlated with the ICU SOFA score suggesting that the levels of Nfl are related to the level of systemic inflammation and organ failure in COVID-pos ICU patients (r = 0.49, p = .003, Pearson). We also performed a correlation analysis of Nfl and results from 33 different clinical blood panel markers. Results from the correlations that were significant (p<.05) are illustrated in **Figure 4B-E**. The log Nfl values were significantly correlated with levels of **(B)** creatinine (r = 0.7, p<.0001, Pearson), (**C)** N terminal pro B type natriuretic peptide (BNP-NT, r = 0.54, p = .008, Pearson), **(D)** prothrombin time (PT) (r = .39, p = .007, Pearson) and **(E)** the white blood cell count (WBC, r = 0.33, p =.006, Pearson). The inserts for each figure show that there are no differences in the levels of each of the blood markers in **Figure 4B-E** between COVID-pos patients with and without neurological complication suggesting the relationship between log Nfl levels and levels of blood creatinine, BNP-NT, PT and WBC were not related to the presence or absence of neurological complications.

**Figure 4.**
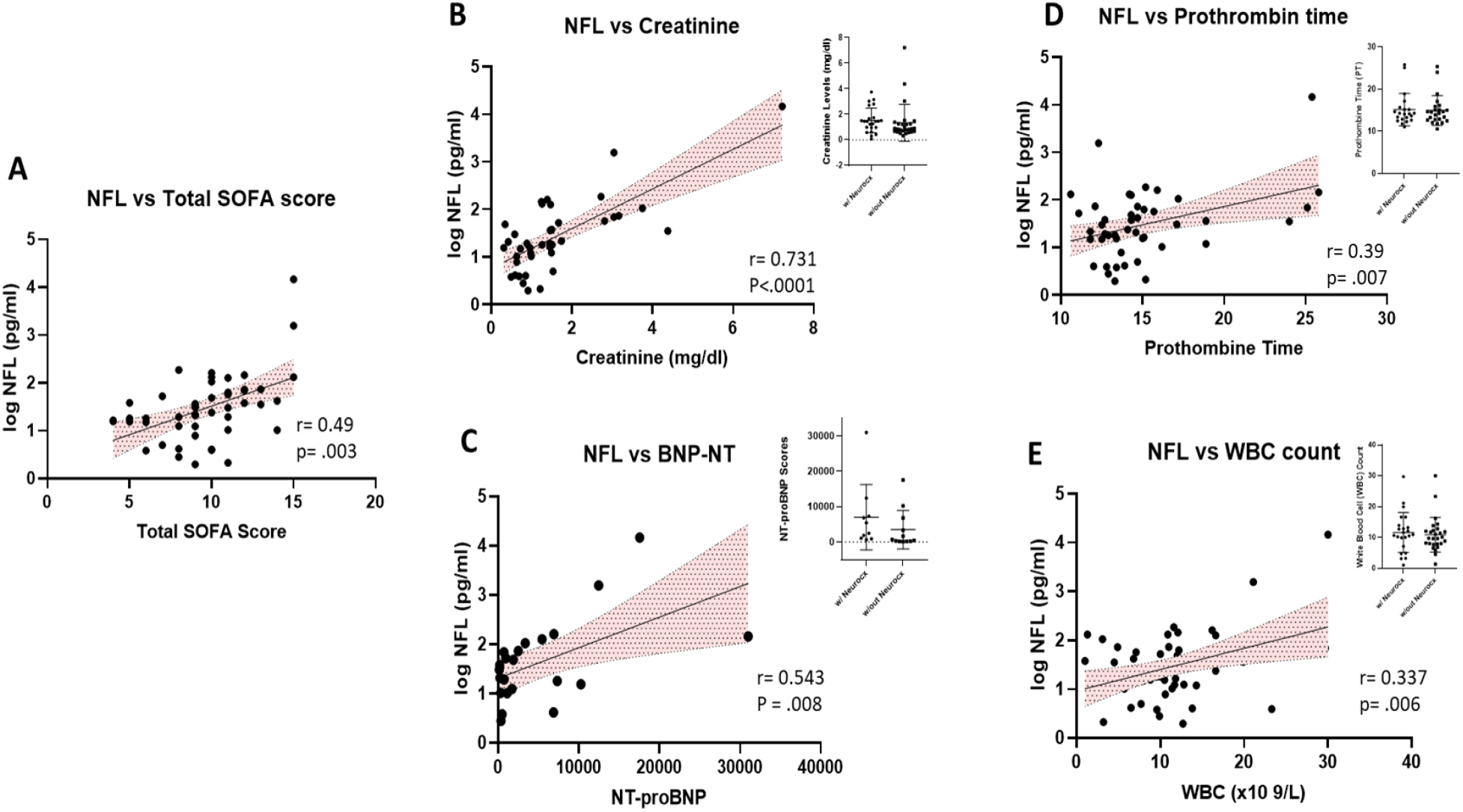
**(A)** Illustrates the positive correlation between the levels of Nfl and the total SOFA score in COVID-pos patients (r = 0.49, p =.003, Pearson). In addition, there was a positive correlation in COVID-pos patients between the levels of Nfl and **(B)** creatinine, r =.73, p<.0001, Pearson), **(C)** BNP-NT, r=0.54, p =.008, Pearson **(D)** prothrombin time, r=0.38, p = .007, Pearson and **(E)** WBC count, r = 0.337, p = .006, Pearson.

## Discussion

In our cohort of 100 patients hospitalized in the ICU in Tucson, Arizona between April 2020 and August 2020, 89 tested positive for COVID-19 and 11 tested negative. We measured serum levels of Nfl in all of these patients and compared them to each other and a cohort of healthy controls (n=8). The mean levels of Nfl in the COVID-pos patients were significantly higher as compared to those observed in COVID-neg patients and healthy controls and suggest that COVID-pos ICU patients may have CNS injury. Of the 50 COVID-pos patients from which we had clinical data, 70% also had at least one or more diagnosis of one of the subtypes of CVD that included heart failure, hypertension (defined as > 140-159/90-99 mm Hg), coronary artery disease, valvular disease, arrhythmia, hyperlipidemia, obesity, and smoking history. The levels of Nfl in these patients with CVD were 205% higher than in those COVID-pos patients those with no CVD. Likewise, those COVID-pos patients with diabetes had 102% higher levels of Nfl than those without diabetes. The levels of Nfl were similar in COVID-pos patients with CVD alone and no neurocx as compared to COVID-pos patients with both CVD and neurocx, suggesting that the increased levels of Nfl in COVID-pos patients with CVD was not dependent on the presence or absence of neurocx.

Numerous studies have shown that Nfl levels in both the serum and CSF have been validated to be able to detect brain injury and axonal damage in individuals with neurodegenerative diseases (12),(13),(14), brain trauma (19) as well as cardiovascular disease and cardiac surgery (17), (18). A number of studies have shown that cognitive impairment and neurodegenerative disease are correlated with vascular disease, inflammation and decreased cerebral brain blood flow (26),(27),(28), (29),(30),(31),. Mechanisms thought to contribute to cognitive impairment in patients with chronic CVD and diabetes include high levels of systemic inflammation (35), altered cerebrovascular autoregulation (38), and microembolism (39). Inflammatory processes play an important role in CVD-related increases in circulating inflammatory mediators are seen in the brain.

An early study of 214 COVID-19 patients in China was among the first to report evidence of neurological complications of SARS-CoV2 infection (5). The symptoms in these patients were categorized into three areas including changes to 1) the central nervous system (CNS) which included patient reports of dizziness, headache, ataxia, impaired consciousness, and acute cerebrovascular disease; 2) the peripheral nervous system (PNS) symptoms (loss of smell, taste and some loss of peripheral sensation); and 3) skeletal muscular dysfunction. In these studies, 41.1% of the subjects exhibited severe COVID-19 disease requiring ventilation. In these patients, 36.3% also had hypertension and 45.5% had neurological manifestations. It has been suggested that the CNS complications in COVID-19 may be due ACE2 expression in the CNS and the PNS (40). Given that ACE2 is known to be expressed in brain endothelial cells within the cerebrovascular system and on glia and neurons (41), it has been suggested that SARS-Cov-2 binding to ACE2 in brain vascular endothelium may result in a compromised blood-brain-barrier (BBB). This compromise would allow entry of the virus into the brain parenchyma leading to virus induced neuronal inflammation and damage (42), (43).

We also found a significant positive correlation of Nfl levels with ICU clinical status and the SOFA score and four blood chemistry measurements including creatinine, prothrombin time (PT), WBC counts and NT-pro-BNP. Alterations in coagulation disorders have recently been reported in other COVID-19 studies (44),(45), (46), (47), (48) and have been suggested to be related to microvascular disease, capillary leakage and poor prognosis in COVID-19 patients. Increases in NT-pro-BNP is a well-known clinical biomarker for heart disease and has also been shown to be affiliated with microvascular disease in the brain, kidney and heart (49) and cardiac complications and mortality rates in COVID-19 patients (50),(51).

Taken together, our data suggest that increased levels of Nfl in COVID-19 ICU patients are related to not only to the neurological complications seen in these patients, but also to the presence and extent of chronic inflammatory diseases such as cardiovascular disease and diabetes. In older adults, chronic systemic inflammatory-related diseases, such as vascular disease, heart failure and hypertension that lead to increased brain and systemic inflammation and decreased brain perfusion, are known to increase the risk for dementia and the development of VCID (52),(53),(54),(55). The COVID-19 related cytokine storm and high levels of proinflammatory cytokines along with hypoxia due to respiratory dysfunction and concomitant CVD seen in COVID-19 ICU patients are likely to result in short and long-term cognitive dysfunction and may accelerate pre-existing cognitive deficits (56), (57), (43).

## Limitations

This study has several limitations. First, there are a limited number of subjects in all groups and an even a further limitation on the numbers of subjects we had access to full clinical data. We were not able to obtain consent from all ICU participants which significantly limited our ability to perform full clinical data comparisons between the COVID-pos and COVID-neg subjects. Also, our healthy control sample, while similar in age-range, was small and included no baseline clinical comorbidity data. Future studies with larger cohorts and complete clinical records will be needed to fully understand the relationship between Nfl and ICU patient status. Lastly, we have no uniform neurological or cognitive assessments in the ICU patients which makes it impossible to correlate these Nfl levels to cognitive function.

## Conclusions

Increased levels of Nfl in COVID-19 ICU patients are correlated with both neurological complications and with the presence of cardiovascular disease and diabetes. Nfl may serve as a biomarker for the risk of cognitive and neurological impairment in COVID-19 patients admitted to the ICU.

## Data Availability

Researchers can request for access to anonymized data from the present study for well-defined research questions that are consistent with the overall research agenda for the cohort. Please contact the corresponding author.

## Acknowledgements

We would like to acknowledge Heidi Erikson and Trina Hughes (Tucson, Arizona) and Anna Pfister and Irina Nilsson (Mölndal, Sweden) for their expert technical assistance.

